# Gait, physical activity, and tibiofemoral cartilage damage: A longitudinal machine learning analysis in the Multicenter Osteoarthritis Study

**DOI:** 10.1101/2022.06.30.22277057

**Authors:** Kerry E. Costello, David T. Felson, S. Reza Jafarzadeh, Ali Guermazi, Frank W. Roemer, Neil A. Segal, Cora E. Lewis, Michael C. Nevitt, Cara L. Lewis, Vijaya B. Kolachalama, Deepak Kumar

## Abstract

**Objective:** To 1) develop and evaluate a machine learning model incorporating gait and physical activity to predict medial tibiofemoral cartilage worsening over two years in individuals without or with early knee osteoarthritis and 2) identify influential predictors in the model and quantify their effect on cartilage worsening.

**Design:** An ensemble machine learning model was developed to predict worsened cartilage MRI Osteoarthritis Knee Score at follow-up from gait, physical activity, clinical and demographic data from the Multicenter Osteoarthritis Study. Model performance was evaluated in repeated cross-validations. The top 10 predictors of the outcome across 100 held-out test sets were identified by a variable importance measure statistic, and their marginal effect on the outcome was quantified by g-computation.

**Results:** Of 947 legs in the analysis, 14% experienced medial cartilage worsening at follow-up. The median (2.5^th^-97.5^th^ percentile) AUC across the 100 held-out test sets was 0.73 (0.65-0.79). Presence of baseline cartilage damage, higher Kellgren-Lawrence grade, greater pain during walking, higher lateral ground reaction force impulse, greater time spent lying, and lower vertical ground reaction force unloading rate were associated with greater risk of cartilage worsening.

**Conclusions:** An ensemble machine learning approach incorporating gait, physical activity, and clinical/demographic features showed good performance for predicting cartilage worsening over two years. While identifying potential intervention targets from the model is challenging, these results suggest that lateral ground reaction force impulse, time spent lying, and vertical ground reaction force unloading rate should be investigated further as potential early intervention targets to reduce medial tibiofemoral cartilage worsening.

**Summary box:** *What are the findings?:*

- Machine learning models predicted cartilage worsening in persons without or with early knee osteoarthritis from gait, physical activity, and clinical and demographic characteristics with a median AUC of 0.73 across 100 held-out test sets.
- High lateral ground reaction force impulse, more time spent lying, and low vertical ground reaction force unloading rate were associated with increased risk of cartilage worsening over two years.

*How might it impact on clinical practice in the future?:*

- Gait and physical activity are some of the only modifiable risk factors for knee osteoarthritis; this study identified three potential intervention targets to slow early knee osteoarthritis progression.

## INTRODUCTION

Knee osteoarthritis (OA) is a progressive, painful joint disease and leading cause of disability, affecting over 350 million adults.(1) While some individuals with advanced disease undergo knee replacement, there is no cure and many experience pain and poor quality of life for decades. Existing structural damage and other risk factors (e.g., obesity, malalignment) can drive further degeneration.(2, 3) Addressing this burden will require early identification of at-risk individuals and discovery of intervention targets that can be addressed before the onset of extensive damage or other risk factors.

Joint loading is one of few modifiable risk factors for knee OA(4) and can be manipulated through gait and physical activity. While prior research has identified gait features associated with medial tibiofemoral knee OA progression,(5) these were typically examined in isolation, in small samples, and/or without accounting for other risk factors. Importantly, little is known about gait and physical activity predictors of progression early in the disease process. Machine learning can identify features in complex datasets that are important to prediction without requiring assumptions about underlying relationships among features, making it useful for exploring gait and physical activity.(6-8)

The Multicenter Osteoarthritis Study (MOST)(9) is a large observational cohort of individuals with and without knee OA where data on gait, physical activity, clinical, and demographic measures are available for machine learning applications. Further, MOST includes MRI exams at multiple timepoints, providing sensitive measures of early joint structural change, including worsening cartilage damage.(10) Using MOST data, our objectives were to (1) build and evaluate a machine learning model to predict medial tibiofemoral cartilage worsening over 2-years from gait, physical activity, clinical, and demographic features in individuals without or with early knee OA, and (2) identify features that contribute most to model prediction and quantify their effect on the outcome.

## METHODS

### Study sample

At 144-months, surviving participants from the original MOST cohort (age 50-79, with or at increased risk for developing knee OA at enrollment) were invited for a return visit. Concurrently, a new cohort (age 45-69, Kellgren-Lawrence grades [KLG] ≤2, with or without knee pain) was enrolled. Participants with inflammatory arthritis or stroke were not included in either cohort. MOST received institutional review board approval from each core site (Boston University, University of Alabama at Birmingham [UAB], University of California San Francisco, University of Iowa [UIowa]). All participants provided informed consent before participating, in accordance with the Helsinki Declaration.

We used data from both cohorts for our baseline (original: 144-month, new: enrollment) and 2-year follow-up (original: 168-month, new: 24-month). MRIs were read for one knee per participant (herein referred to as the “study knee”) at baseline and 2-years. If both knees had readable baseline and follow-up images, the knee with better quality images was read. We excluded participants with KLG >2 in the study knee to focus on early disease (Figure 1). We excluded participants with history of knee or hip replacement (either leg), steroid or hyaluronic acid injection during the past 6 months (either knee), or regular use of walking aids. Finally, we excluded participants who did not undergo MRI assessment or with gait or physical activity data quality issues (described later).

**Figure 1.**
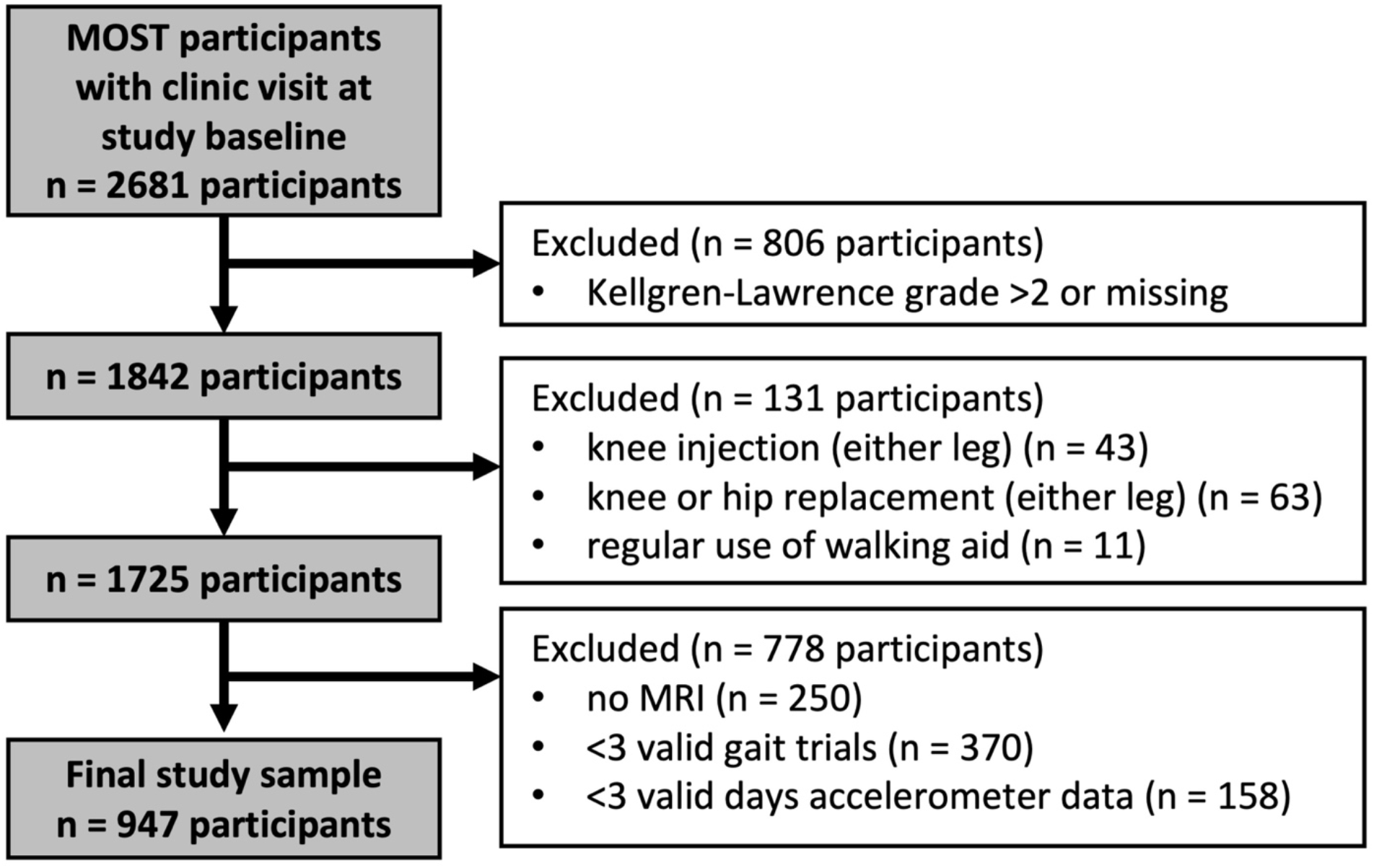
Study sample from the Multicenter Osteoarthritis Study

### Patient and public involvement

Currently, patients and the public are not involved in the design, conduct, reporting, or dissemination plans for research projects utilizing MOST data.

### Equity, diversity, and inclusion statement

The authors include women and men with training in engineering and various clinical specialties from Asia, Europe, and North America. This study included participants (58.2% women) from two North American clinical sites with various self-reported racial identities (Table 1). Sex, site, and race were accounted for in our analyses (described later); however, we did not examine socioeconomic status.

**Table 1.**
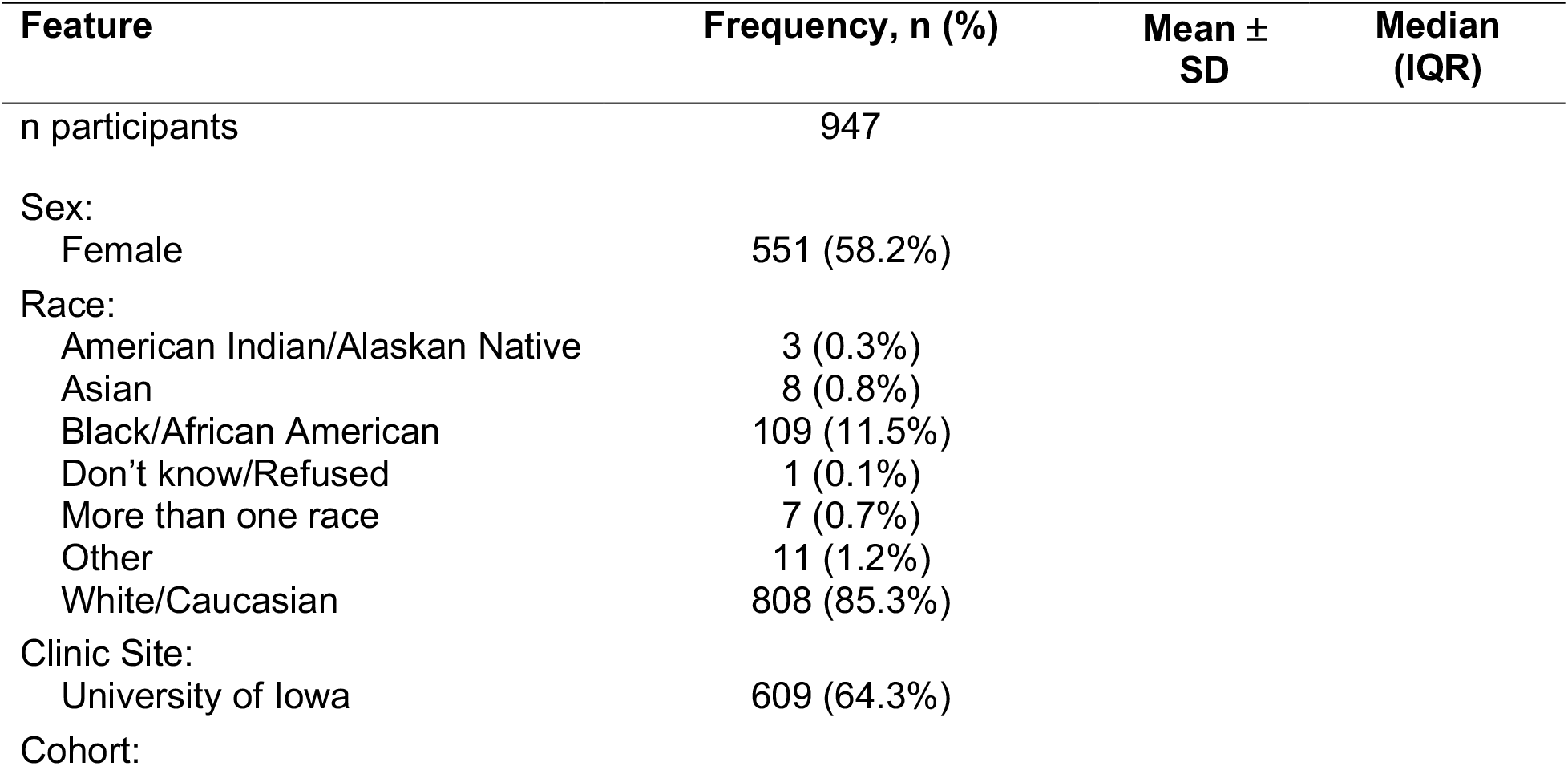
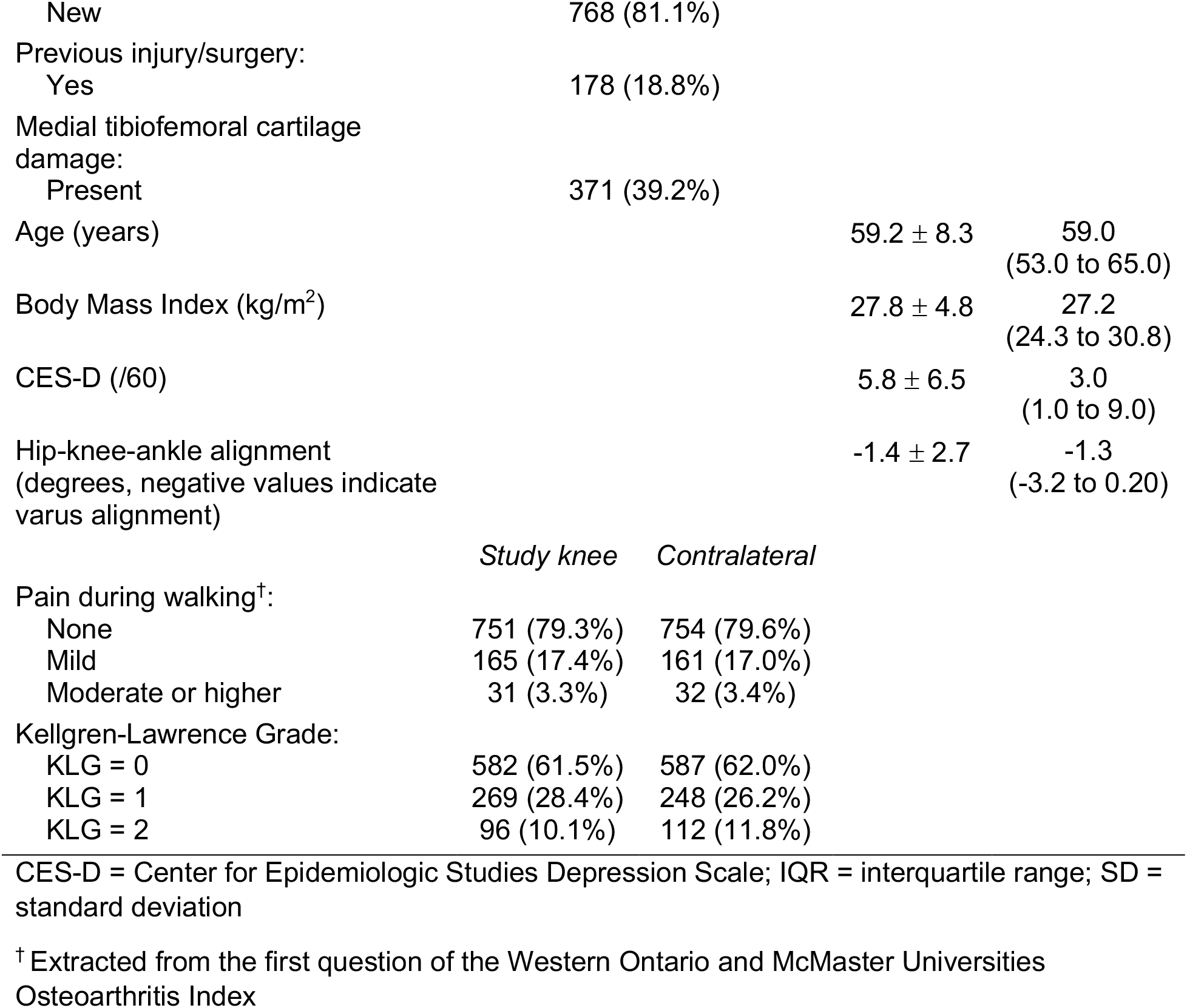
Baseline demographics and clinical characteristics

### Exposures

#### Clinical and demographic features

Model inputs included clinical and demographic factors (11-18) that are both independent risk factors for OA and affect gait/physical activity (i.e., confounders based on hypothesized directed acyclic graphs (19)). Sex, age, body mass index (BMI), race, clinic site, and prior history of knee injury or surgery were recorded at baseline. Given small samples in multiple categories of race (Table 1), particularly at UIowa, race and site were combined into a single feature with 3-levels: UAB non-White (n=117), UAB White (n=221), UIowa (n=609). Participants completed the Western Ontario and McMaster Universities Osteoarthritis Index (WOMAC)(20) and Center for Epidemiologic Studies Depression Scale (CES-D), and had posterior-anterior and lateral weight bearing radiographs taken, which were read for KLG (21). Hip-knee-ankle alignment was read from baseline long-limb radiographs for the new cohort and long-limb radiographs taken at the 60-month visit for the original cohort. Pain during walking was extracted from the first question of WOMAC (categorized as ‘no,’ ‘mild,’ or ‘moderate or higher’).

#### Gait features

Three-dimensional (3D) ground reaction force (GRF) data were recorded (1000 Hz) while participants walked at self-selected speed across a portable force platform embedded in a 5.3-meter walkway (AccuGait, AMTI Inc., Watertown, MA, USA). At least five trials were acquired per leg, with the first excluded as an acclimatization trial. Legs with ≤3 remaining trials where the foot landed completely on the force plate were retained for analysis. For each trial, we extracted commonly used 3D GRF metrics (Figure 2), “toe-out” angle defined by Chang et al.,(22) stance time, and walking speed. We normalized all timing features to stance phase (i.e., % stance). GRFs were not amplitude-normalized given the inclusion of BMI in the model and to avoid issues with interpreting ratios.(23) We averaged each feature across trials for each leg.

**Figure 2.**
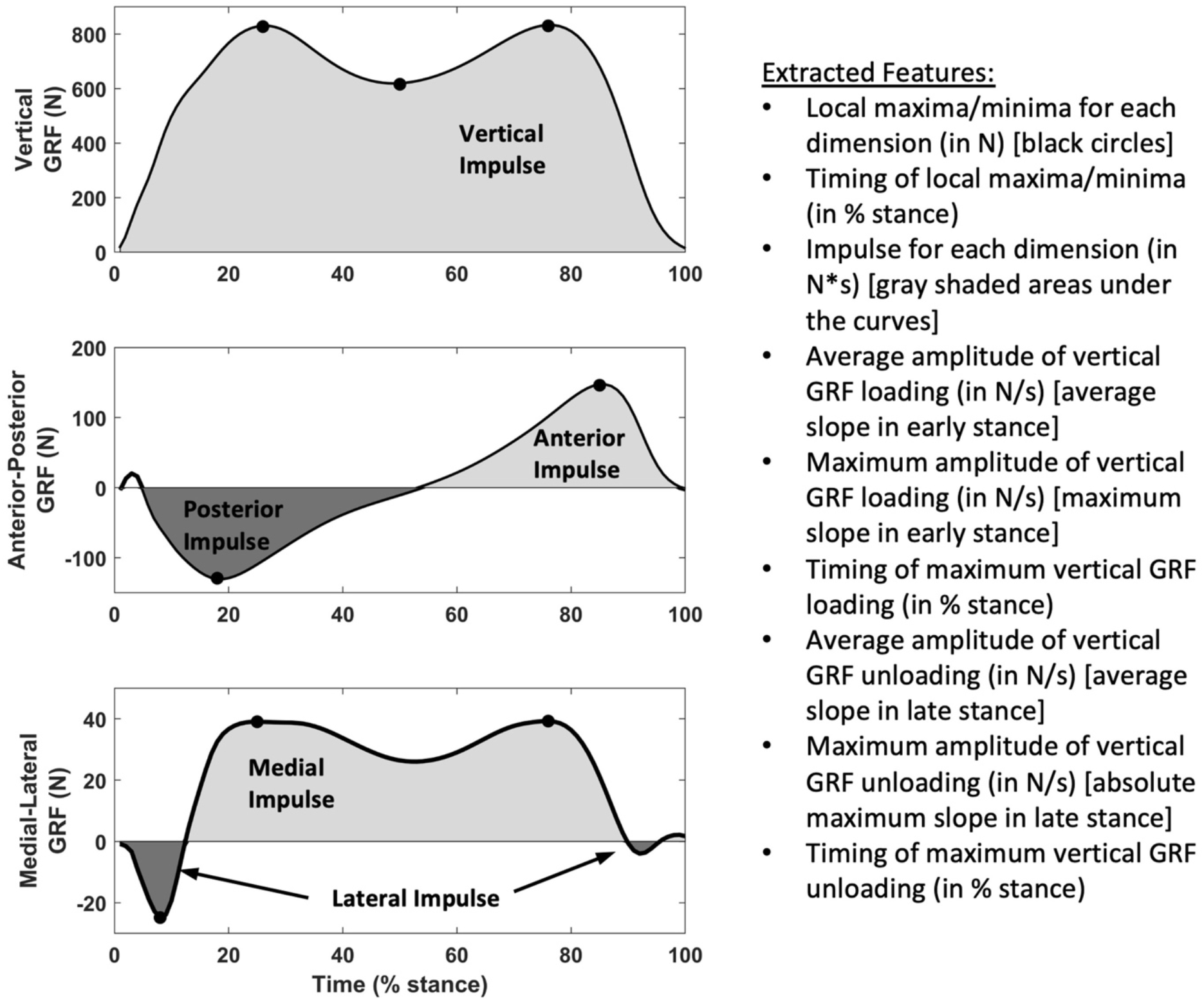
Features extracted from ground reaction force (GRF) data

#### Physical activity features

Participants wore an activity monitor (AX3, Axivity, Newcastle upon Tyne, UK) consisting of a tri-axial accelerometer and temperature sensor on the lower back (centered over the midpoint of L5-S1) for 7 days at baseline, with 3D acceleration sampled at 100 Hz with a range of ±8g. Non-wear was defined as periods ≥10 minutes with no movement and verified using the temperature sensor.(24) Data for each axis were bandpass filtered (0.2-20 Hz, 4^th^ order Butterworth filter). Summary metrics were calculated for each day: step count, time spent walking, time spent lying, and mean 3D signal vector magnitude (overall magnitude of acceleration across all dimensions, Equation 1). Time spent walking and lying were expressed as % wear time to account for differences in wear time among individuals.(25) Metrics were averaged across all valid days (defined as ≥10 hours of wear time/day (26)). We excluded participants with <3 valid days.(27)

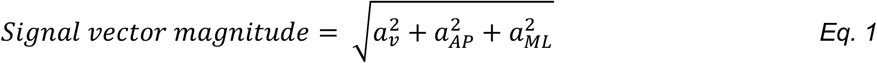

### Outcome

Two musculoskeletal radiologists (AG, FWR) scored the severity of cartilage damage in 5 medial tibiofemoral subregions of the study knee at each timepoint using the MRI Osteoarthritis Knee Score (MOAKS).(28) We defined medial cartilage worsening as any increase in area and/or depth in at least one of the 5 subregions over the 2-year period, as done previously.(10, 29)

### Machine learning model

Model development was performed in R (v4.2.2). We examined Spearman correlations between all continuous features and for near perfect correlations (ρ>0.85), selected one feature to retain for analysis (Table 2 shows retained gait and physical activity features). We used the predictive mean matching algorithm within the multiple imputation by chained equations (MICE) framework (v3.13.0) to impute missing exposure data (<0.1% dataset).(30) We randomly split the data into 70% train and 30% test, maintaining the same proportion of outcome in both datasets.(31) Continuous features were scaled and centered to have zero mean and unit variance.

**Table 2.** Baseline gait and physical activity features

| Feature | Mean ± SD | Median (IQR) |
| --- | --- | --- |
| **GRF impulses (N\*s):** |  |  |
| Vertical GRF impulse | 454.5 ± 113.7 | 439.5<br>(370.5 to 523.5) |
| Medial GRF impulse | 18.9 ± 8.1 | 17.6<br>(13.0 to 23.1) |
| Lateral GRF impulse | 1.6 ± 1.0 | 1.5<br>(0.9 to 2.1) |
| Anterior GRF impulse | 23.9 ± 7.0 | 23.1<br>(18.5 to 28.3) |
| **GRF local maxima (N):** |  |  |
| Vertical GRF 1<sup>st</sup> peak | 850.6 ± 179.5 | 826.9<br>(718.2 to 962.9) |
| Posterior GRF peak | 134.1 ± 41.1 | 128.5<br>(103.5 to 156.4) |
| Early medial GRF peak | 47.6 ± 15.8 | 45.7<br>(36.3 to 57.5) |
| **GRF loading and unloading rates (N/s):** |  |  |
| Maximum instantaneous vertical GRF loading rate | 11555 ± 4077 | 10981<br>(8776 to 13379) |
| Maximum instantaneous vertical GRF unloading rate | 10204 ± 2289 | 9983<br>(8564 to 11536) |
| **Timing of GRF local maxima/minima (% stance):** |  |  |
| Vertical GRF 1<sup>st</sup> peak | 25.9 ± 3.5 | 25.7<br>(23.8 to 28.0) |
| Vertical GRF 2<sup>nd</sup> peak | 75.5 ± 2.5 | 75.9<br>(74.5 to 77.0) |
| Vertical GRF valley (midstance minimum) | 48.3 ± 3.5 | 48.3<br>(46.0 to 50.8) |
| Posterior GRF peak | 17.5 ± 2.4 | 17.5<br>(16.1 to 18.9) |
| Anterior GRF peak | 85.4 ± 1.4 | 85.5<br>(84.6 to 86.4) |
| Early lateral GRF peak | 7.1 ± 1.7 | 7.2<br>(6.1 to 8.2) |
| Early medial GRF peak | 26.3 ± 5.7 | 26.0<br>(22.2 to 30.1) |
| Late medial GRF peak | 72.3 ± 5.7 | 73.5<br>(70.0 to 76.3) |
| **Spatiotemporal parameters:** |  |  |
| Gait speed (m/s) | 1.35 ± 0.2 | 1.34<br>(1.21 to 1.48) |
| Stance time (s) | 0.7 ± 0.1 | 0.7<br>(0.7 to 0.8) |
| Angle formed by center of pressure path and direction of travel, "toe-out angle" (degrees, negative values indicate varus) | -3.7 ± 5.3 | -3.5<br>(-7.2 to -0.2) |
| **Accelerometer derived physical activity measures:** |  |  |
| Step count | 9554 ± 3919 | 9149<br>(6812 to 11533) |
| Time spent lying (% total wear time) | 9.2 ± 10.4 | 5.6<br>(2.0 to 12.5) |
| Mean signal vector magnitude (milligravity) | 4.0 ± 1.4 | 3.8<br>(3.1 to 4.7) |

GRF = ground reaction force; IQR = interquartile range; SD = standard deviation

Our goal was to predict the binary cartilage worsening outcome from baseline GRF, accelerometer, and clinical/demographic data. We used “super learning” (v1.4.2),(32) an ensemble machine learning approach that combines several candidate algorithms to enhance prediction accuracy above and beyond individual algorithms (Figure 3). We selected candidate learners to include diverse learning strategies while being computationally feasible, as recommended by Phillips et al.(33) Using the training dataset, candidate learners were trained through 5-fold cross validation. Corresponding predictions on out-of-fold samples were used to develop a meta learner that optimized the weight (i.e., contribution) of each individual learner. We then applied this model to the held-out test set to assess its performance by area under the receiver operating characteristic curve (AUC) and mean square error (MSE).

**Figure 3.**
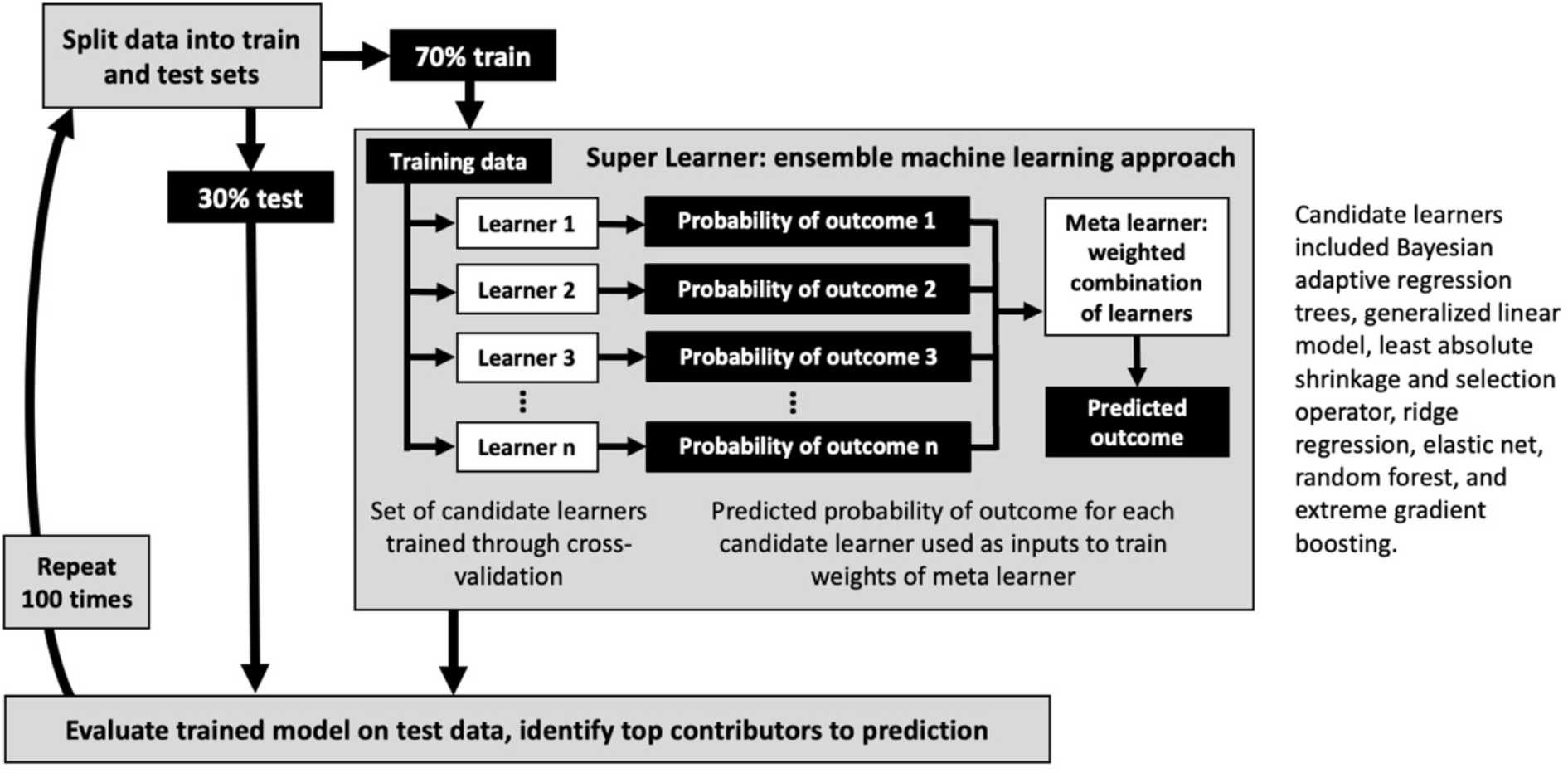
Machine learning model development and evaluation

To test robustness and reproducibility of the model training and testing, we used repeated cross-validation, i.e., repeated the process of randomly splitting the data into train and test, training the super learner, and evaluating its performance on the held-out test set. Here we report median (i.e., 50-th percentile), 2.5- and 97.5-th percentile AUC and MSE across 100 iterations.

### Identification of influential predictors

To assess the contribution of each feature to model prediction, for each of the 100 iterations, 35 additional models were trained on the training set (each excluding one of the features included in the full model). These models were applied to the test set and a Variable Importance Measure (VIM) statistic was calculated for each feature for each iteration based on the size of the risk difference between the full model and the model fit without the feature. Thus, 35 VIMs were produced per iteration. The top contributors to prediction for each iteration were identified as the 10 features with the highest VIMs. We defined “influential predictors” as the 10 features that most frequently appeared as top contributors across the 100 iterations.

### Marginal causal risk differences

To quantify the effect of influential predictors identified from the super learner model on cartilage worsening, we utilized parametric g-computation.(34) Continuous variables were quantized into tertiles. For each predictor, we calculated the marginal causal risk difference of each category of the predictor on cartilage worsening, compared to the corresponding reference category, using riskCommunicator (v 1.0.0); 95% confidence intervals were calculated using 1000 bootstrap samples.(35) Different risk factors may be associated with OA initiation versus progression; thus, we explored sensitivity analyses stratified by baseline cartilage damage (i.e., lesion in ≥1 subregion). Only 6% of knees without baseline damage had cartilage worsening at follow-up, thus we focused our sensitivity analysis on those with baseline damage (Supplement).

## RESULTS

### Model performance

Of 947 participants, 133 (14%) experienced cartilage worsening in the study knee over 2-years. Across 100 iterations, the median (2.5- and 97.5-th percentiles) AUC and MSE on the held-out test sets were 0.73 (0.65-0.79) and 0.11 (0.09-0.13), respectively.

### Influential predictors

The features most frequently appearing as top contributors to prediction across 100 iterations (and frequency of appearance) were baseline medial tibiofemoral cartilage damage (100), KLG (98), lateral GRF impulse (46), pain during walking (45), time spent lying (35), timing of the vertical GRF 1^st^ peak (31), vertical GRF impulse (30), early medial GRF peak (29), timing of the vertical GRF 2^nd^ peak (28), and the maximum instantaneous vertical GRF unloading rate (28).

### Marginal risk differences

Marginal risk differences from the g-computation analyses (Figure 4) can be interpreted as the difference in risk of cartilage worsening per 100 individuals in the given category compared to the referent category. Presence of cartilage damage, higher KLG, greater lateral GRF impulse, greater pain during walking, greater time spent lying, and lower vertical GRF unloading rate at baseline were associated with increased risk of cartilage worsening (Figure 4). Point estimates were similar in the sensitivity analysis (Supplement).

**Figure 4.**
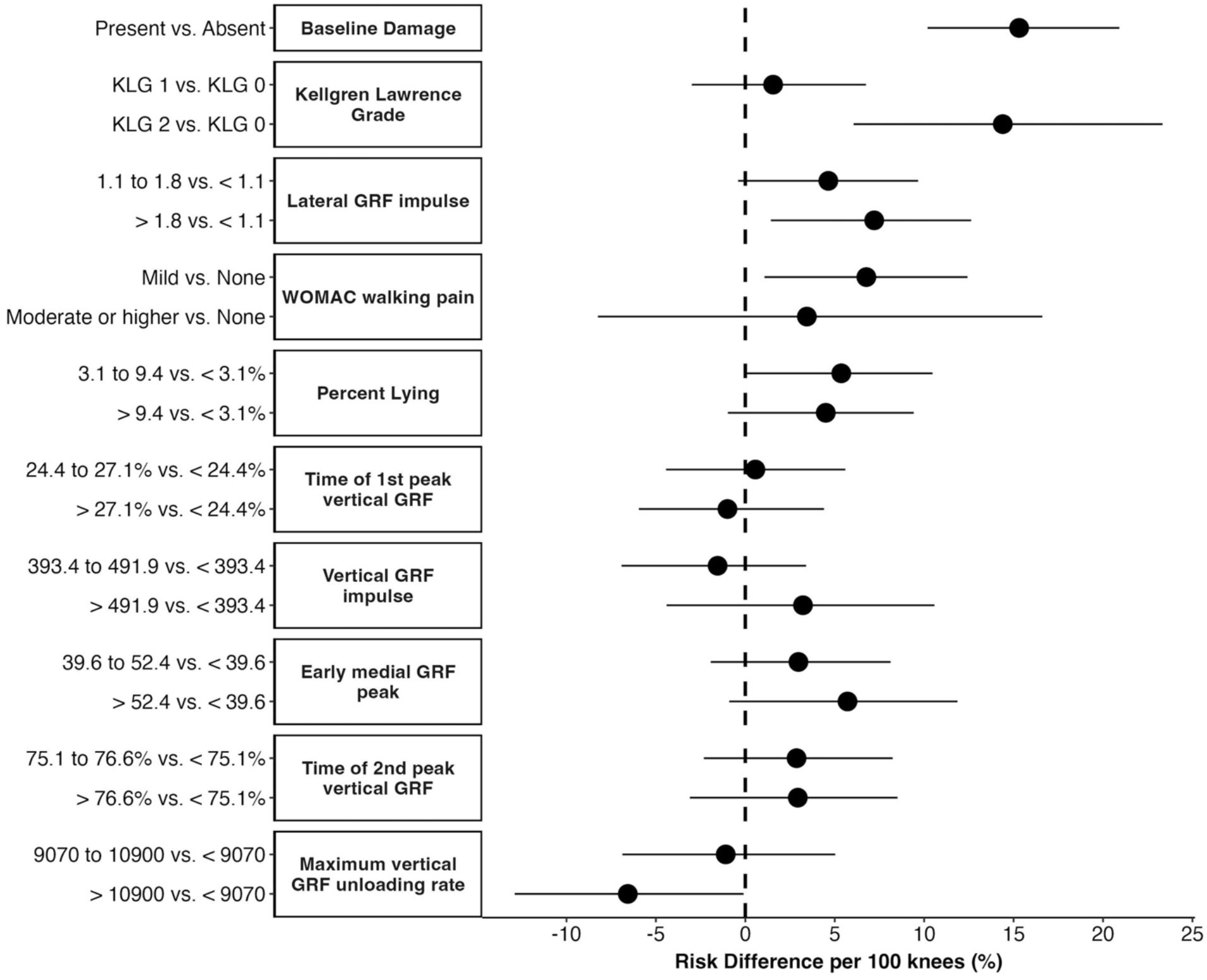
Causal risk differences for influential predictors identified from the machine learning model

## DISCUSSION

An ensemble machine learning approach incorporating baseline gait, physical activity, and clinical/demographic features showed good performance predicting medial tibiofemoral cartilage worsening over 2-years in knees with KLG ≤2. While determining the relationships among predictors and outcomes in machine learning models is challenging, our analysis suggests that high lateral GRF impulse, high time spent lying, and low vertical GRF unloading should be investigated further as potential targets to reduce cartilage worsening.

### Model performance

Our model performance is comparable to other machine learning models predicting OA progression from clinical/demographic data. Du et al. reported AUCs of 0.70-0.79 for predicting radiographic worsening (increase in KLG, medial, or lateral joint space narrowing) over 2-years from baseline cartilage damage MRI features in those with KLG 0 to 4.(36) Tiulpin et al. reported AUCs of 0.73-0.75 for predicting worsening (increase in KLG or knee joint replacement) over 7-years from baseline age, sex, BMI, injury, surgery, WOMAC, and KLG in individuals with KLG < 2.(37) The current model achieved similar AUC for predicting cartilage worsening over 2-years in individuals with KLG ≤ 2, with the added benefit of providing information about potentially modifiable gait and physical activity predictors.

Prior longitudinal gait studies typically examined knee-specific loading (e.g., knee adduction moment) rather than GRFs, often in samples of 15 to 300 knees.(5) Correspondingly, few addressed clinical/demographic confounders, incorporated physical activity, or examined performance in held-out test sets. Further, many were conducted in samples with established OA (KLG ≥2), limiting their potential to identify at-risk individuals early in the disease process or identify early intervention targets. Our sample included 947 individuals with KLG ≤2, who predominantly had no or mild pain during walking, and thus were younger with lower BMI than previously reported samples (mean age 59.2 vs. 62.0 years, BMI 27.8 vs. 29.2 kg/m^2^).(18)

### Predictors of OA progression

The super learner identified multiple influential gait and physical activity predictors of cartilage worsening. Of these, the g-computation analyses found baseline lateral GRF impulse, time spent lying, and vertical GRF unloading rate were associated with cartilage worsening. The 7.2% estimate of risk difference for lateral GRF impulse suggests that for every 100 individuals in the highest tertile, there are 7.2 individuals who experience cartilage worsening who would not experience worsening in they were in the lowest tertile. Accordingly, approximately 14 (i.e., 1/0.072) individuals with lateral GRF impulse at the highest tertile would be needed to observe an increase in the number of individuals with cartilage worsening by 1 person. In a cross-sectional study in the same cohort, we previously reported that limbs with radiographic OA, with or without knee pain, have higher lateral GRFs in early stance compared to limbs without radiographic OA or pain.(38) The current results suggest lateral GRF may also play a role in progression. The 5.4% increased risk of cartilage worsening for the middle versus lowest tertile of time spent lying, along with prior research showing greater sedentary time is associated with future functional decline (39) and lower quality of life(40), suggests reducing sedentary time should be investigated as a potential intervention target. The physiological reason for the 6.6% decreased risk of cartilage worsening in the highest versus lowest tertile of vertical GRF unloading rate is not clear and warrants further exploration.

The appearance of structure and symptom features as influential predictors is not surprising given that these are established risk factors. Of note, despite only 10.1% of the sample having established radiographic OA (KLG=2), 39.2% had baseline cartilage damage, and both appeared as influential predictors in the model. The g-computation analysis identified a 15.3% increased risk of cartilage worsening for every 100 individuals with baseline damage compared to no damage, and a 14.4% increased risk for KLG 2 versus 0. The lack of difference for KLG 1 versus 0 may highlight limitations of the KLG scoring system, which does not reflect tissue-level damage well, particularly in early disease.(41, 42) Along with these structural measures, knees with mild pain had an increased risk of cartilage worsening (6.8%) compared to those with no pain. The large confidence interval for moderate and higher pain could stem from the small proportion of knees (3% of sample) and/or heterogeneity in this category.

### Clinical implications

The utility of this model for risk screening is debatable, as it requires GRF data. While faster to collect than joint moments, collecting GRFs requires specialized equipment (force platform). Future advances in wearable technologies may facilitate gait data capture during everyday life, including estimates of GRFs,(43, 44) improving the potential of this type of model as a risk screening tool.

This model identified potential gait and physical activity intervention targets for further study. Interestingly, two influential predictors (baseline damage, KLG) appeared as top contributors in ≥98% of the iterations while others appeared less consistently (<50%). While we removed highly correlated features, this may result in part from predictors that capture similar constructs (e.g., four features collectively describing an important construct could each appear 25% of the time). Similarly, our g-computation approach provides insight into causal pathways but does not account for concurrent changes in several risk factors. An important motivation for using machine learning was to address potential interactions among predictors. While it is challenging to identify these relationships from the model, the lack of consistency in top contributors could indicate a need for simultaneous intervention on several features rather than a single feature, opening interesting avenues for future study.

### Strengths and limitations

Strengths include the large sample, investigation of gait and physical activity in early disease, use of machine learning to address inter-related predictors, and use of g-computation to quantify their effects. These strengths expand existing literature by accounting for demographics and clinical characteristics, examining multiple gait and physical activity features, and testing the model on held-out data. Our sample was largely White with little to no pain during walking; these results may not generalize to diverse populations or those with severe symptoms. Lateral or patellofemoral worsening could have been present in both outcome groups, causing noise in the model. While we adjusted for a diverse set of confounders, as in any observational study there may be residual unmeasured confounding. Knee loading (e.g., knee adduction moment) may provide additional predictive power but kinematics are not available in MOST, limiting comparison to prior gait studies and insight into mechanisms by which features such as lateral GRF impulse affect structure. We are unaware of other large datasets with gait, physical activity, and MR outcomes that could be used for external validation, however we assessed reproducibility with repeated cross-validation. Better characterization of dynamic physical activity patterns may also improve model performance and identification of relevant intervention targets.

## Conclusion

Using an ensemble machine learning approach, we predicted medial tibiofemoral cartilage worsening over 2-years in persons without and with early radiographic osteoarthritis with good performance on held-out samples. Additionally, we identified baseline gait and physical activity measures associated with cartilage worsening that may be potential early intervention targets, including lateral GRF impulse, time spent lying, and vertical GRF unloading rate.

## Data Availability

Data are available in a public, open access repository. Data from the Multicenter Osteoarthritis Study are available through the National Institute on Aging Research Biobank: https://agingresearchbiobank.nia.nih.gov/. Public use datasets from the 144- and 168-month visits of the Multicenter Osteoarthritis Study will be available at this website in 2023.

## ETHICS STATEMENT

### Patient consent for publication

Not applicable.

### Ethics approval

Ethical approval for the Multicenter Osteoarthritis Study was obtained at each of the four core sites (Boston University, University of Alabama at Birmingham, University of California San Francisco, University of Iowa) and all participants provided informed consent prior to participating.

## ACKNOWLEDGEMENTS

The authors would like to acknowledge the contributions of the MOST participants and clinic staff.

## Twitter

@kecostello, @srjafarz, @ali_guermazi, @FrankRoemer1, @ProfCaraLewis, @vkola_lab, @ProfDeepakKumar

## Contributors

Responsibility for the integrity of the work as a whole: K.E. Costello, D. Kumar

Conception and design: K.E. Costello, D. Kumar, D.T. Felson

Analysis and interpretation of the data: All authors

Drafting of the article: K.E. Costello

Critical revision of the article for important intellectual content: All authors

Final approval of the article: All authors

Provision of study materials or patients: D.T. Felson, C.E. Lewis, M.D. Nevitt, N.A. Segal

