## Supplement for "Gait, physical activity, and tibiofemoral cartilage damage: A longitudinal machine learning analysis in the Multicenter Osteoarthritis Study"

**Sensitivity analysis examining subsample with baseline cartilage damage**

In the subsample of knees with baseline cartilage damage (Table S1), 26% had cartilage worsening at 2-year follow-up. For each predictor, we calculated the marginal causal risk difference of each category of the predictor on cartilage worsening, compared to the corresponding reference category using g-computation. Continuous variables were categorized using tertile cutpoints calculated from the full sample (as detailed in the main manuscript). The models included the same predictors as in the main manuscript except for baseline cartilage damage (i.e., 9 total predictors included).

As in the main analysis, in the subsample with baseline cartilage damage, the g-computation analysis identified an increased risk of cartilage worsening for individuals with KLG 2 versus 0 (17.9% per 100 individuals) and for pain during walking of mild versus none (16.3% per 100 individuals) (Figure S1). In the main analysis a lateral ground reaction force (GRF) impulse of 1.8 N*s or higher compared to <1.1 N*s had a higher risk of cartilage worsening. In the subsample, the point estimate was similar to the main analysis (6.1% versus 7.2% per 100 individuals), but the 95% confidence interval included zero. Similarly, point estimates were similar for the middle versus lowest tertile of time spent lying (7.1% versus 5.4% per 100 individuals) and for the highest versus lowest tertile of maximum vertical GRF unloading rate (7.8% versus 6.6%) but both 95% confidence intervals included zero. These wider confidence intervals could be related to the smaller sample size of this subsample or heterogeneity within the subsample.

Table S1. Baseline demographics and clinical characteristics for subsample with baseline cartilage damage

| **Feature** | **Frequency, n (%)** | | **Mean ± SD** |
|---|---|---|---|
| n participants | 371 | | |
| Sex: | | | |
| Female | 183 (49.3%) | | |
| Race: | | | |
| American Indian or Alaskan Native | 1 (0.3%) | | |
| Asian | 3 (0.8%) | | |
| Black or African American | 36 (9.7%) | | |
| Don’t know/Refused | 0 (0.0%) | | |
| More than one race | 3 (0.8%) | | |
| Other | 3 (0.8%) | | |
| White or Caucasian | 325 (87.6%) | | |
| Clinic Site: | | | |
| University of Iowa | 254 (66.0%) | | |
| Cohort: | | | |
| New | 268 (72.2%) | | |
| Previous injury/surgery: | | | |
| Yes | 89 (32.8%) | | |
| Age (years) | | | 61.2 ± 8.6 |
| Body Mass Index (kg/m$^2$) | | | 28.2 ± 4.8 |
| Center for Epidemiologic Studies Depression score (/60) | | | 5.3 ± 5.4 |
| Hip-knee-ankle alignment (degrees, negative values indicate varus alignment) | | | -1.9 ± 2.7 |
| | *Study knee* | *Contralateral* | |
| WOMAC pain during walking: | | | |
| None | 293 (79.0%) | 294 (79.2%) | |
| Mild | 62 (16.7%) | 62 (16.7%) | |
| Moderate or higher | 16 (4.3%) | 15 (4.0%) | |
| Kellgren-Lawrence Grade (KLG): | | | |
| KLG = 0 | 156 (42.0%) | 171 (46.1%) | |
| KLG = 1 | 136 (36.7%) | 130 (35.0%) | |
| KLG = 2 | 79 (21.3%) | 70 (18.9%) | |

SD = standard deviation; WOMAC = Western Ontario and McMaster Universities Osteoarthritis Index

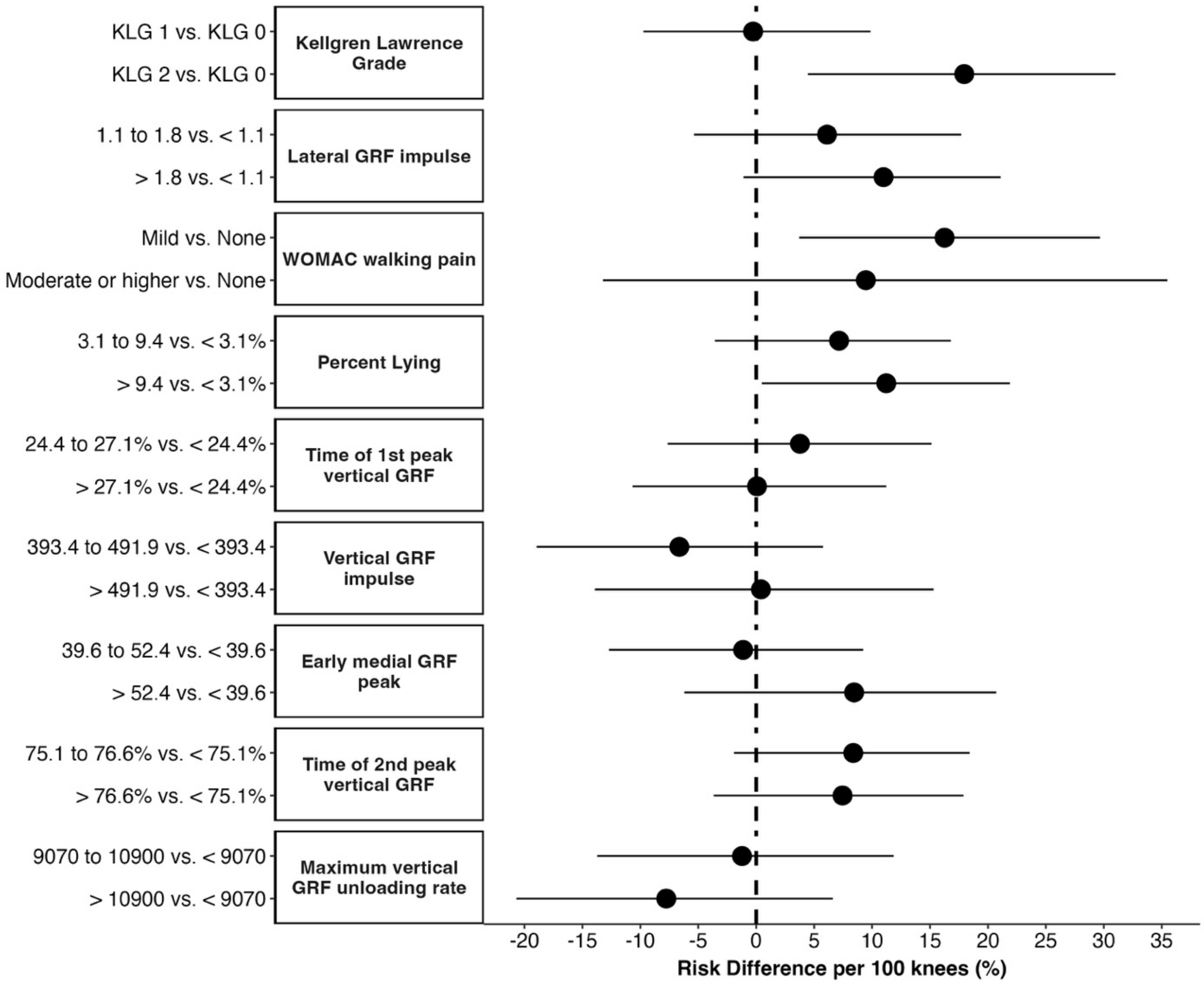


Figure S1. Causal risk differences in the subsample with baseline cartilage damage for influential predictors identified from the machine learning model
